# A tale of two variants: Spread of SARS-CoV-2 variants Alpha in Geneva, Switzerland, and Beta in South Africa

**DOI:** 10.1101/2021.06.10.21258468

**Authors:** Christian L. Althaus, Stephanie Baggio, Martina L. Reichmuth, Emma B. Hodcroft, Julien Riou, Richard A. Neher, Frédérique Jacquerioz, Hervé Spechbach, Julien Salamun, Pauline Vetter, Carolyn Williamson, Nei-yuan Hsiao, Wolfgang Preiser, Mary-Ann Davies, Richard J. Lessells, Tulio de Olivera, Laurent Kaiser, Isabella Eckerle

## Abstract

Several SARS-CoV-2 variants of concern (VOC) are spreading rapidly in different regions of the world. The underlying mechanisms behind their transmission advantage remain unclear. We measured viral load in 950 individuals and found that infections with variant Alpha exhibit a higher viral load and longer viral shedding compared to non-VOC. We then used a transmission model to analyze the spread of variant Alpha in Geneva, Switzerland, and variant Beta in South Africa. We estimated that Alpha is either associated with a 37% (95% compatibility interval, CI: 25–63%) increase in transmissibility or a 51% (95% CI: 32–80%) increase of the infectious duration, or a combination of the two mechanisms. Assuming 50% immune evasion for Beta, we estimated a 23% (95% CI: 10–37%) increase in transmissibility or a 38% (95% CI: 15–78%) increase of the infectious duration for this variant. Beta is expected to outgrow Alpha in regions where the level of naturally acquired immunity from previously circulating variants exceeds 20% to 40%. Close monitoring of Alpha and Beta in regions with different levels of immunity will help to anticipate the global spread of these and future variants.

## INTRODUCTION

Novel SARS-CoV-2 variants emerged independently in different geographic regions of the world. According to the definition of the World Health Organization (WHO) as of 25 February 2021, a SARS-CoV-2 variant is defined as a variant of concern (VOC) if it has been demonstrated to be associated with an increase in transmissibility or profound changes in the epidemiology, increased virulence or change in clinical disease presentation; or decrease in effectiveness of public health and social measures or available diagnostics, vaccines, and therapeutics (World Health Organization, 2021). To date, there are four variants identified that fulfill the criteria, namely variant Alpha, first detected in the UK, variant Beta, first detected in South Africa, variant Gamma that emerged in Brazil, and variant Delta, first documented in India. All four variants have become the dominant circulating virus in the affected regions within a short time period, raising concerns about their increased fitness and transmission advantage (Abdool Karim and de Oliveira, 2021).

For Alpha, several studies have estimated an increased transmissibility between 40% to 100% in the United Kingdom, the United States, Denmark, and Switzerland (Leung et al., 2021; Davies et al., 2021a; Volz et al., 2021; Chen et al., 2021). A higher transmissibility per contact with an infectious person is supported by findings that patients infected with Alpha (or S-gene target failure, SGTF) appear to have lower Ct values (Davies et al., 2021b) and higher viral loads (Ratcliff et al., 2021). A recent study compared 1453 Alpha cases with 977 non-Alpha cases and found a 1.0 higher mean log_10_ viral load and a 2.6 times higher cell culture replication probability in Alpha cases (Jones et al., 2021). However, others found no differences in viral burden for SGTF (Walker et al., 2021). In addition, preliminary data suggest that Alpha could also be associated with extended periods of viral shedding when compared to previously circulating variants (Kissler et al., 2021). In contrast, evasion from naturally acquired immunity seems to play little to no role for the transmission advantage of Alpha as neutralization by both convalescent sera from previous infections, as well as vaccine-derived antibodies, were able to neutralize Alpha in a similar or only slightly reduced manner (Abdool Karim and de Oliveira, 2021).

The mechanisms of the transmission advantage of Beta are less well understood. While an increased transmissibility and/or a longer infectious duration cannot be ruled out (Pearson et al., 2021), the constellation of mutations in the spike receptor-binding-domain (particularly mutation E484K and K417N) and the N-terminal domain has been associated with escape from monoclonal antibodies (mAb) and polyclonal serum mediated neutralization (Tegally et al., 2021; Wibmer et al., 2021; Cele et al., 2021; Collier et al., 2021). Gamma, with which Beta shares E484K and other critical mutations, was estimated to evade 21–46% of protective immunity elicited by previous infection with non-VOC (Faria et al., 2021). In order to anticipate the global spread of Alpha, Beta and other variants, it is critically important to understand the consequences of these altered transmission characteristics in different epidemiological settings.

In this study, we aimed at better understanding the mechanisms that result in a transmission advantage of Alpha and Beta. First, we measured viral load in 950 individuals infected with either Alpha or non-VOC. Second, we analyzed the increase in the proportion of Alpha in Geneva, Switzerland, and Beta in South Africa using a transmission model. We then estimated the fitness advantage of the two variants considering the following mechanisms: i) increase in transmissibility, ii) increase of infectious duration, and iii) immune evasion. We compared the fitness advantage of both variants at different levels of naturally acquired immunity, and discussed the implications of our findings for anticipating the further spread of these and other SARS-CoV-2 variants.

## METHODS

### Data

#### Viral load

We assessed individuals presenting at the outpatient SARS-CoV-2 screening site at the Geneva University Hospitals presenting for routine diagnostic SARS-CoV-2 testing. The majority of patients had symptoms compatible with SARS-CoV-2 infection and a small proportion were asymptomatic contacts. All participants were ≥ 16 years old with suspected SARS-CoV-2 infection according to the local governmental testing criteria, i.e., suggestive symptoms for coronavirus disease 2019 (COVID-19) and/or recent exposure to a SARS-CoV-2 positive person. We included only nasopharyngeal swabs (NPS) collected from symptomatic patients with known date of symptom onset. We analyzed all NPS samples using the Cobas® SARS-CoV-2 RT-PCR assay on the 6800 system (Roche), targeting the E and ORF1 gene, or the TaqPath assay (Thermofisher), targeting the N, ORF1, and S gene. To convert Ct values into SARS-CoV-2 RNA copy numbers/ml, we performed serial testing of dilutions of cultured SARS-CoV-2, which were quantified by using *in vitro* transcribed RNA obtained from the European Virus Archive by using the Charité E gene assay (Corman et al., 2020; Baggio et al., 2020).

From 13 January 2021 to 24 March 2021, we re-screened all positive samples with a diagnostic Ct value ≤ 32 with a single nucleotide polymorphism (SNP) specific RT-PCRs for mutations 501Y and E484K, allowing to assess presence or absence of the mutation by melting curve analysis (VirSNiP SARS Spike 501Y, VirSNiP SARS Spike E484K, TibMolBiol, Berlin). To identify samples belonging to Alpha, we defined presence of the 501Y mutation and absence of the E484K mutation. Next generation sequencing of a subset of positive specimens confirmed that this combination of mutations correlated with Alpha. In order to increase our sample size for the comparison group for non-VOC viruses, we also included patient samples from the same setting tested from 1 October 2020 to 16 December 2020, a time when no VOCs were circulating in Geneva or Switzerland. We excluded all asymptomatic cases, patients with missing values for date of symptom onset, symptom onset > 12 days, other types of material, and a Ct value of initial diagnostic RT-PCR > 32.

#### Viral variants

To track the spread of Alpha in Geneva, Switzerland, we relied on the identification of Alpha described above. To cover the period of November and December 2020, we used sequence data from randomly chosen samples from Geneva that were submitted to GISAID by the Swiss SARS-CoV-2 Sequencing Consortium. The data on the proportion of Alpha in Geneva, Switzerland, have been made available on the following website: https://ispmbern.github.io/covid-19/variants/. For Beta in South Africa, we retrieved all South African SARS-CoV-2 sequences from the GISAID database as of 20 January 2021 (*n* = 2986, collected from 6 March 2020 to 6 January 2021) (Shu and McCauley, 2017). We excluded three sequences with unknown collection date, leaving 2983 sequences for analysis.

### Model

Competitive spread between variant and non-variant (‘wild-type’) strains of SARS-CoV-2 can be described within the susceptible-infected-recovered (SIR) framework by the following two ordinary differential equations:

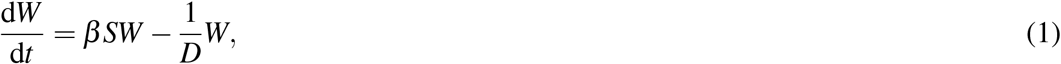

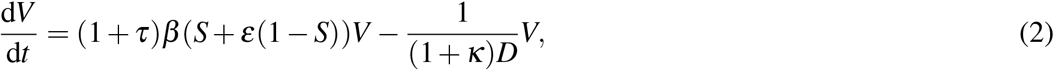

where *W* and *V* are individuals infected with wild-type and variant, respectively, and *S* the population of susceptibles. *β* is the transmission rate and *D* the infectious duration of the wild-type. The fitness advantage of the variant can act via three different mechanisms:

1. *Increase in transmissibility*: The transmission rate of the variant is increased by the factor *τ*.
2. *Increase of infectious duration*: The infectious duration of the variant is increased by the factor *κ*.
3. *Immune evasion*: The variant can partially evade the acquired immunity from previous infections by the wild-type (1 − *S*). Immune evasion can vary from complete cross-protection (*ε* = 0) to full evasion (*ε* = 1).

One can show that the proportion of the variant among all infections increases according to logistic growth (Marée et al., 2000):

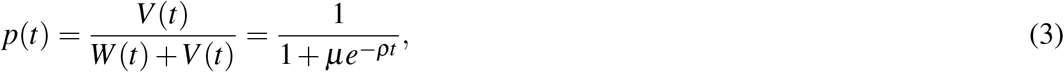

where *µ* = *W* (0)*/V* (0) and *ρ* corresponds to the difference in the net growth rates between the variant and the wild-type:

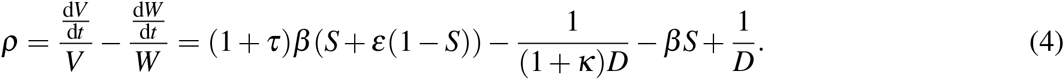

Eq. 4 can be solved algebraically for *τ, κ* or *ε*. If the transmission advantage acts via a single mechanism only, we obtain the following simplified solutions. First, the increased transmissibility of the variant is given by *τ* = *ρ/*(*β S*), assuming there is no change in the infectious duration and no immune evasion (*κ* = *ε* = 0). Since the effective reproduction number of the wild-type is *R*_*w*_ = *βSD*, we obtain *τ* = *ρD/R*_*w*_. Second, the increased infectious duration of the variant is given by *κ* = *ρ/*(1*/D* − *ρ*), assuming there is no change in transmissibility and no immune evasion (*τ* = *ε* = 0). Finally, assuming there is no change in transmissibility nor the infectious duration (*τ* = *κ* = 0), the level of immune evasion is given by *ε* = *ρ/*(*β* Ω) = *ρD*(1 − Ω)*/*(Ω*R*_*w*_), where Ω = 1 − *S* corresponds to the proportion of the population with previously acquired immunity against earlier variants, i.e., the cumulative incidence or seroprevalence, at the time the variant starts to grow.

We estimated *ρ* by fitting a logistic growth model (binomial regression) to the proportion *p*(*t*) of Alpha in Switzerland, Geneva, and Beta in South Africa. To propagate the uncertainty, we constructed 95% compatibility intervals (CIs) for *τ, κ* and *ε* from 10,000 parameter samples of *ρ*, the generation time *D*, the effective reproduction number of the wild-type *R*_*w*_, and the seroprevalence Ω (Amrhein et al., 2019). We assumed a normally distributed generation time with a mean of 5.2 days and a standard deviation of 0.8 days (Figure 1A) (Ganyani et al., 2020). We sampled from publicly available estimates of the daily effective reproduction number based on confirmed cases during the early growth phase of the variants in Geneva, Switzerland, (1 November 2020 to 31 January 2021; range: 0.58–1.04) and South Africa (1 September 2020 to 31 October 2020; range: 0.90–1.12) (https://github.com/covid-19-Re) (Figure 1B) (Huisman et al., 2020). In Geneva, Switzerland, seroprevalence was estimated at 21.1% (95% credible interval: 19.2–23.1%; *n* = 4,000) in samples collected from 23 November 2020 to 23 December 2020 (Figure 1C) (Stringhini et al., 2021). In South Africa, seroprevalence was estimated at 30.2% (95% CI: 28.8–31.2%; *n* = 4,387) in samples collected from 17 August 2020 to 25 November 2020 (Shinde et al., 2021). We ignored vaccine-induced immunity as vaccination uptake was still low during the study periods in both countries. All data and R code files are available on GitHub: https://github.com/calthaus/sarscov2-variants.

**Figure 1.**
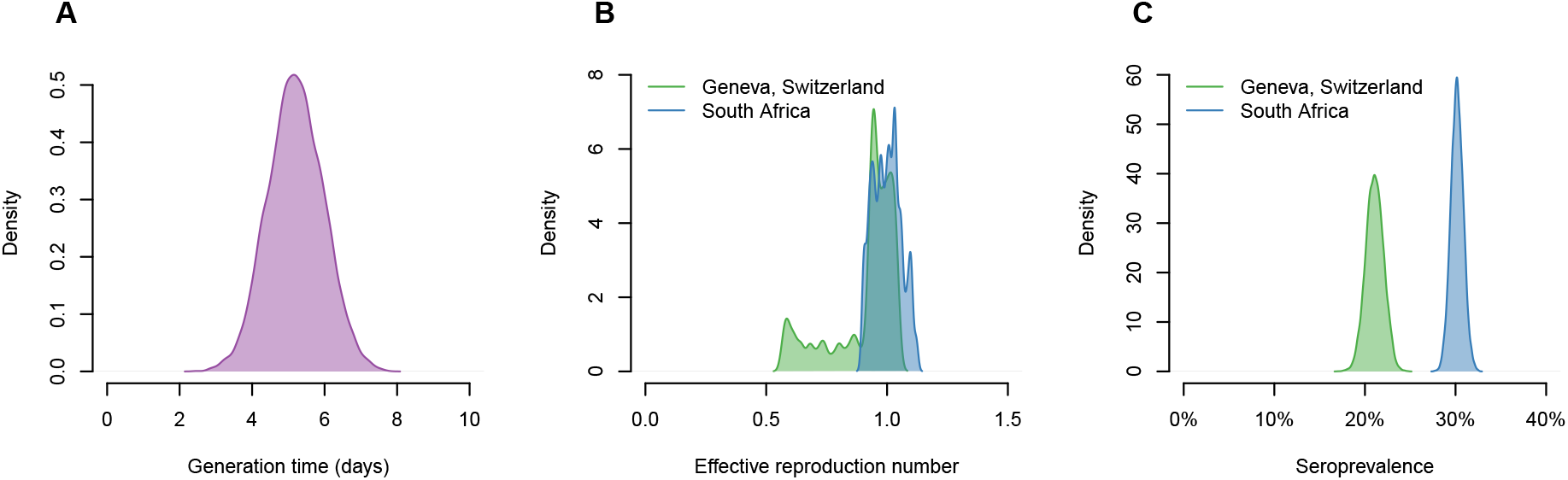
Parameter distributions of the generation time, effective reproduction number, and seroprevalence. A: Generation time based on Ganyani et al. (2020). B: Effective reproduction number during the early growth phase of the variants (https://github.com/covid-19-Re) (Huisman et al., 2020). C: Seroprevalence estimates for Geneva, Switzerland (Stringhini et al., 2021), and South Africa (Shinde et al., 2021).

## RESULTS

We analyzed viral load in a total of 950 specimens from Geneva, Switzerland (604 non-VOC, 346 Alpha). We found a higher mean viral load for Alpha compared to non-VOC (7.4 vs. 6.9 SARS-CoV-2 log10 RNA copies/ml, *p* < 0.001) (Figure 2A). Analyzing viral load by day post onset of symptoms showed a delayed decrease in viral load for Alpha compared to non-VOC (Figure 2B). Notably, viral load for non-VOC fell below the threshold for presence of infectious (culturable) virus (10^6^ SARS-CoV-2 RNA copies/ml) at day 6 to 11. In contrast, viral load remained above that threshold for B.1.1.7 in samples taken from day 6 to 11 post onset of symptoms. Together, these data suggest that Alpha exhibits a transmission advantage that is mediated by either an increased transmissibility or a longer infectious duration, or a combination of both mechanisms.

**Figure 2.**
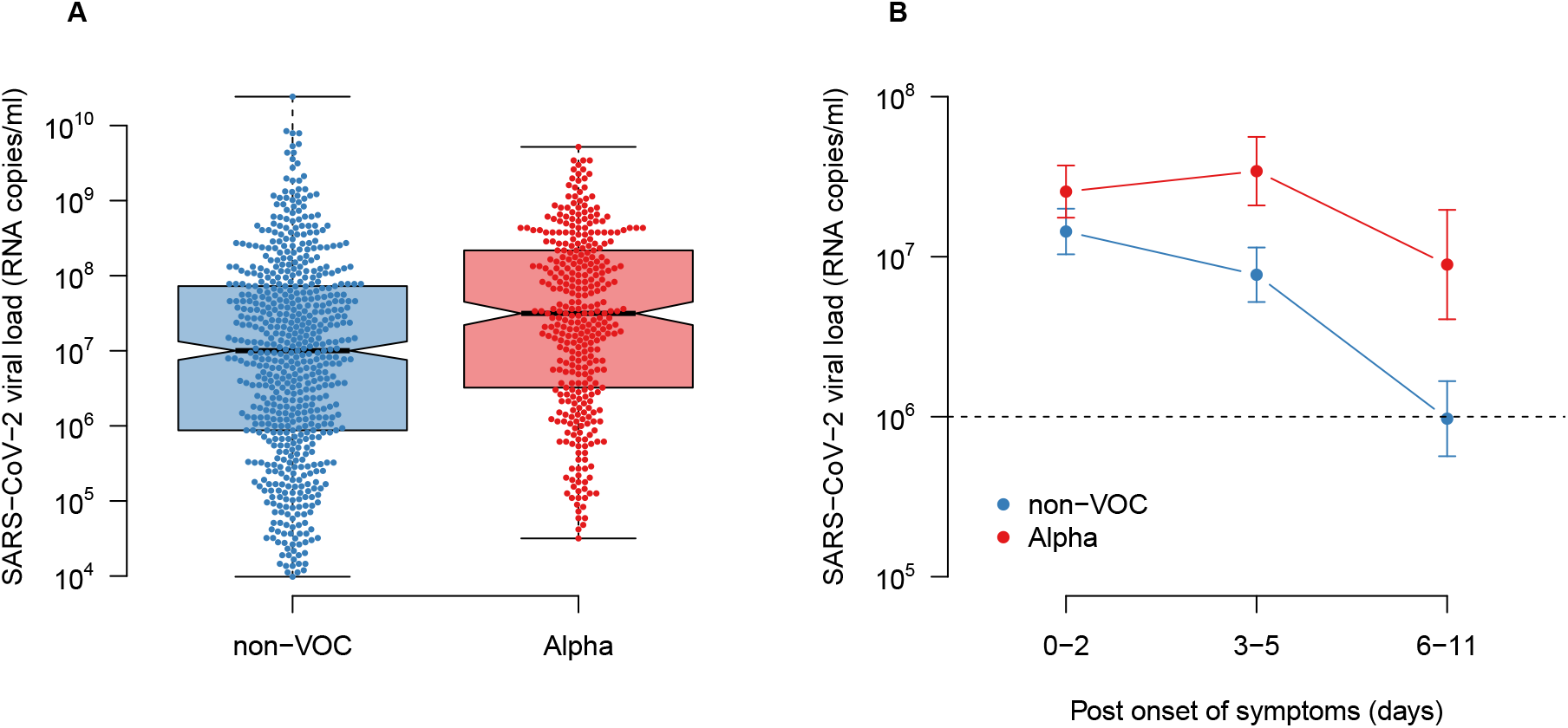
SARS-CoV-2 viral load for non-VOC and Alpha. A: Comparison of overall viral load. B: Comparison of viral load per day post onset of symptoms. Data: 950 (604 non-VOC, 346 Alpha) individuals from the outpatient screening site at the Geneva University Hospitals, Switzerland. Error bars correspond to the 95% compatibility intervals of the mean. Dashed line: Assumed threshold for presence of infectious virus (10^6^ SARS-CoV-2 RNA copies/ml).

Alpha was first detected in Geneva, Switzerland, in a sample from 22 December 2020 and almost completely replaced the previously circulating variants by the end of March 2021 (Figure 3A). We estimated the logistic growth rate of the proportion of Alpha at 0.065 (95% CI: 0.060– 0.071) per day. This corresponds to either a 37% (95% CI: 25–63%) increase in transmissibility or a 51% (95% CI: 32–80%) increase of the infectious duration. As expected, immune evasion alone can be ruled out as an explanation for the observed spread of Alpha as seroprevalence levels in Geneva were not high enough even if evasion was complete (94% of parameter samples resulted in *ε* > 1) (Figures 4B and C).

**Figure 3.**
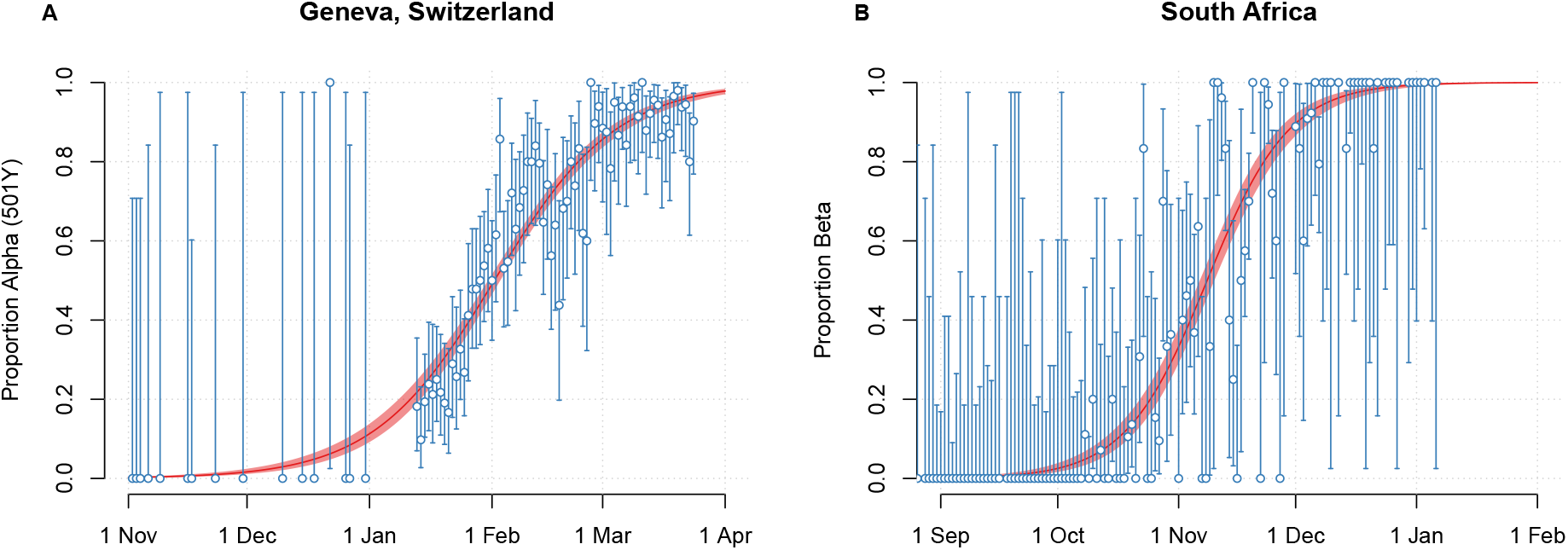
Increase in the proportion of SARS-CoV-2 variants among confirmed cases. A: Alpha in Switzerland, Geneva. B: Beta in South Africa. Error bars and shaded areas correspond to 95% compatibility intervals of the data (blue) and model (red), respectively.

**Figure 4.**
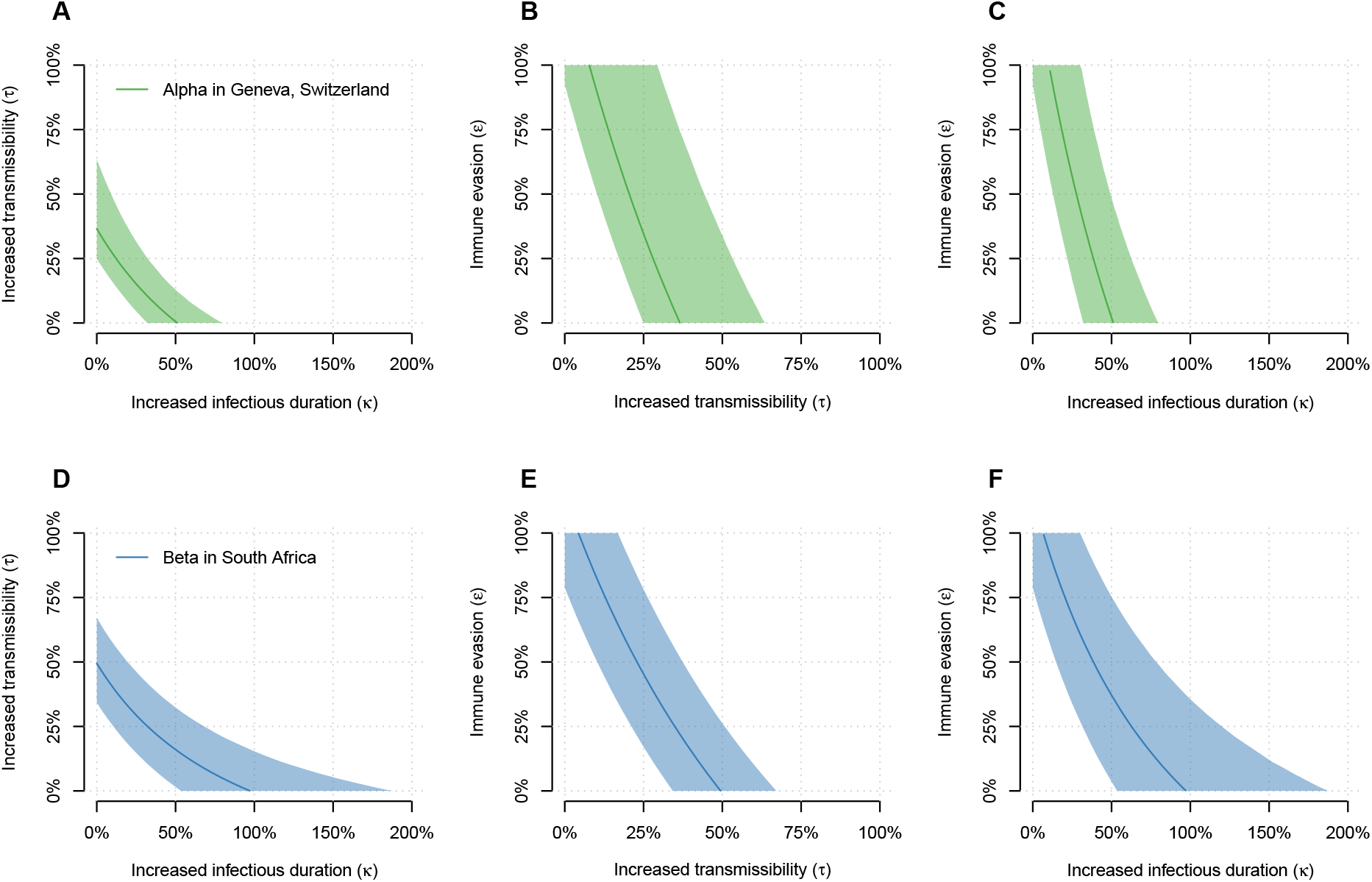
Relationship between increase in transmissibility, increase of infectious duration, and immune evasion. Top row: Alpha in Geneva, Switzerland. Bottom row: Beta in South Africa. Lines and shaded areas correspond to the median and 95% compatibility intervals.

Beta was first detected in South Africa in a sample from 8 October 2020 and practically replaced all previously circulating variants by the end of December 2020 (Figure 3B). We estimated the logistic growth rate of the proportion of Beta at 0.095 (95% CI: 0.085–0.106) per day. This corresponds to either a 49% (95% CI: 34–67%) increase in transmissibility or a 97% (95% CI: 54–187%) increase of the infectious duration. Complete immune evasion alone would require a seroprevalence level of 33% (95% CI: 25–40%) to explain the spread of B.1.351, which is only slightly higher than the estimated level of 30% in South Africa. Nevertheless, since 78% of parameter samples resulted in *ε* > 1 (Figures 4E and F), we conclude that Beta is likely to be associated with an increased transmissibility and/or an increased infectious duration in addition to partial immune evasion.

Based on our analysis, it is not possible to quantify the degree of immune evasion for Beta (Figures 4E and F). An analysis of Gamma – which shares E484K with Beta – in Brazil estimates that this variant evades 25–61% of protective immunity elicited by previous infection with other variants (Faria et al., 2021). Assuming 50% immune evasion for Beta, we estimated an additional 23% (95% CI: 10–37%) increase in transmissibility or a 38% (95% CI: 15–78%) increase of the infectious duration, which is less than for Alpha without immune evasion (Figures 4E and F).

In regions where both Alpha and Beta are present, the existing level of protective immunity against previously circulating variants, i.e., the cumulative incidence or seroprevalence, may influence which variant will outgrow the other, although transmission heterogeneity and other epidemic drivers will mediate this. We estimated the expected growth advantage of both variants as a function of seroprevalence, assuming no immune evasion for Alpha and varying levels of immune evasion for Beta (Figure 5). Depending on the level of immune evasion, Beta is expected to outgrow Alpha when the level of naturally acquired immunity against previously circulating variants exceeds 20% to 40%.

**Figure 5.**
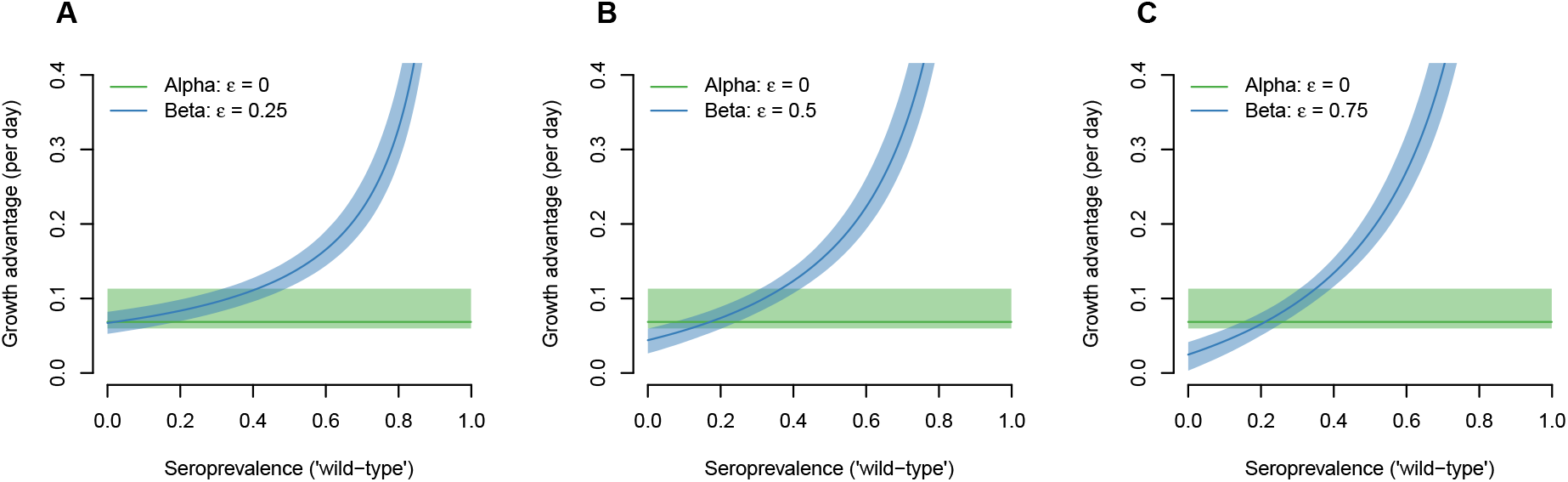
Growth advantage of Alpha and Beta over ‘wild-type’ variants of SARS-CoV-2. For Alpha, we assumed no immune evasion (*ε* = 0), i.e., the growth advantage is constant and independent of the seroprevalence. For Beta, we assumed immune evasion of 25% (A), 50% (B), and 75% (C). The growth advantage is relative to a ‘wild-type’ variant with *R*_*w*_ = 1. Lines and shaded areas correspond to the median and 95% compatibility intervals.

## DISCUSSION

We used clinical and epidemiological data to better understand the mechanisms and implications of the transmission advantage of the SARS-CoV-2 variants Alpha and Beta. We found that Alpha infections exhibit a higher viral load and longer viral shedding compared to non-VOC. Using a transmission model, we estimated that Alpha is either associated with a 37% (95% CI: 25–63%) increase in transmissibility or a 51% (95% CI: 32–80%) increase of the infectious duration. Assuming that Beta results in partial immune evasion, we estimated that Beta exhibits a lower increase in transmissibility and/or the infectious duration compared to Alpha. Nevertheless, our analysis suggests that Beta might be expected to outgrow Alpha in regions where the level of naturally acquired immunity against previously circulating variants exceeds 20% to 40%.

A strength of our study is the combination of clinical and epidemiological data to analyze the mechanisms and the epidemiological implications of the transmission advantage of Alpha and Beta. The transmission model allowed us to study the competitive spread between variant and non-variant strains of SARS-CoV-2 within the framework of evolutionary and population biology. We considered multiple uncertainties, such as the underlying epidemic dynamics and the level of population immunity.

Our study has a number of limitations. First, we inferred specimens from Geneva, Switzerland, as being Alpha by using mutation specific RT-PCR only. However, NGS surveillance of circulating variants in Switzerland confirmed that presence of 501Y was closely correlated with Alpha. Second, we only included samples with a Ct value ≤ 32 to account for testing selection for mutation specific RT-PCR, so absolute viral loads could be biased towards higher values. Third, the threshold for infectious virus (10^6^ SARS-CoV-2 RNA copies/ml) was assessed for non-VOC at the beginning of the pandemic (Wölfel et al., 2020; L’Huillier et al., 2020). As no experimental data on this threshold have been published for Alpha, we assumed that Alpha shows a similar pattern for the presence of culturable virus compared to non-VOC. Fourth, the transmission model did not allow us to quantify the individual contribution of the different mechanisms to the transmission advantage. Fifth, we assumed an exponentially distributed generation time for estimating the increase in transmissibility. A delta distributed generation time would result in slightly higher estimates (Davies et al., 2021a; Volz et al., 2021; Chen et al., 2021). Sixth, estimates of the effective reproduction number based on confirmed cases come with considerable uncertainty. We took this uncertainty into account by sampling from daily estimates over a period of 2 to 3 months. Finally, we used seroprevalence estimates from single studies (South Africa not being based on population representative sampling) and did not consider an increase of seroprevalence during the early growth phase of the variants, heterogeneity across the population, or a potential waning of antibodies.

Our findings support the notion that the transmission advantage of Alpha is likely to be mediated via a higher viral load (increase in transmissibility) and/or longer viral shedding (increase of infectious duration). The estimated increase in transmissibility for B.1.1.7 is in good agreement with the lower end of earlier estimates (Leung et al., 2021; Davies et al., 2021a; Volz et al., 2021; Chen et al., 2021). In addition, we also estimated a potential increase of the infectious duration, i.e., the generation time. These findings have important implications for infection control. A higher transmissibility per contact requires an additional reduction in contacts to prevent further spread. In contrast, the transmission advantage of a longer infectious duration could be compensated by early case finding and isolation, particularly in low incidence settings with efficient contact tracing capacities. The typically used isolation period also appears to be sufficient for Alpha, as no considerable increase in SARS-CoV-2 infections among healthcare workers has been reported.

Immune evasion is arguably of bigger concern than increases in transmissibility or the infectious duration, especially if there is similar evasion of vaccine-elicited immunity. Similar to Gamma, Beta appears to exhibit partial immune evasion. The finding that Beta is not expected to outgrow Alpha in regions where the cumulative incidence of infections with previously circulating variants does not exceed 20% is in agreement with the observation that Beta does not seem to replace Alpha in Switzerland or Denmark (https://cevo-public.github.io/Quantification-of-the-spread-of-a-SARS-CoV-2-variant, https://www.covid19genomics.dk). Similarly, it has been shown that Gamma does not seem to be able to outcompete Alpha in Italy (Stefanelli et al., 2021). On the other hand, Alpha might not be able to outcompete Beta in South Africa. Hence, Beta might outgrow Alpha in some areas of Europe that experience a high cumulative incidence of SARS-CoV-2 in 2020.

Our study helps to understand the consequences of the altered transmission characteristics of Alpha and Beta in different epidemiological settings. The presented modeling framework, which considers three different mechanisms that result in a transmission advantage of VOCs, can be readily applied to data sets from other regions and countries. More research is needed to better understand how VOCs affect symptomaticity and disease severity, and how they respond to cellular and humoral immune responses elicited by natural infection and vaccines. We conclude that the further spread of Alpha and Beta will strongly depend on the level of acquired immunity from previously circulating variants. Hence, it will be important to closely monitor the spread of these and other VOCs in regions with varying levels of naturally acquired immunity and vaccination uptake.

## Data Availability

All data and R code files are available on GitHub: https://github.com/calthaus/sarscov2-variants.

## Acknowledgments

We would like to thank the Network for Genomic Surveillance in South Africa (NGS-SA) which includes the National Health Laboratory Service, the National Institute for Communicable Diseases, the University of Cape Town, Stellenbosch University, the University of Free State, and the KwaZulu-Natal Research Innovation & Sequencing Platform at the University of KwaZulu-Natal.

